# A prospective, split-Face, comparative study of combined Q-switch Nd:YAG laser and microdermabrasion versus Q-switch Nd:YAG laser alone in treatment of melasma

**DOI:** 10.1101/2021.05.19.21257291

**Authors:** Parvaneh Hatami, Hamed Nicknam Asl, Zeinab Aryanian

**Author notes:** **Correspondence to** Zeinab Aryanian, Autoimmune Bullous Diseases Research Center, Razi hospital, Tehran University of Medical Sciences, Tehran, Iran.

## Abstract

**Background:** Melasma is a common disorder of hyperpigmentation with a great effect on quality of life. In an uncontrolled study, a novel combination treatment approach, using a low-fluence Q-switched (QS) Neodymium-doped Yttrium Aluminium Garnet (Nd:YAG) Laser in conjunction with microdermabrasion was shown to be effective in clearance of melasma(1).

**Objective:** The purpose of the current prospective, controlled study was to determine if microdermabrasion, when combined with Nd:YAG laser, produced significant improvement of melasma compared to Nd:YAG laser alone.

**Methods:** Twenty patients with bilaterally distributed facial melasma were subjected to QS-Nd:YAG laser therapy with microdermabrasion on one side and laser therapy alone on the other side of their faces every four weeks for three sessions. Photographs were obtained before treatment, one month and three months postoperatively.

**Results:** Although both sides improved from baseline to the 24th week, there was no statistically significant difference between both sides in regards to MASI (Melasma Area and Severity Index) as well as PGA (Patient Global melasma improvement Assessment) scores.

**Conclusion:** Combination therapy with Q-switched Nd:YAG laser and microdermabrasion seems to be as effective as Q-switched Nd:YAG laser alone and doesn’t have any advantage or disadvantage in comparison with conventional laser therapy.

## Introduction

Melasma is a common cutaneous disorder characterized with symmetrical brown macules and patches with indistinct borders, typically on cheek, forehead and upper lip (2). Melasma can involve all skin types, but it is known to be more prevalent in females of darker skin types (Fitzpatrick skin types IV to VI), especially East Asians (2,3).This pigmentation disorder is often a significant source of distress to those affected and has considerable effects on patient’s quality of life as a matter of beauty(4).

The etiology of this common disorder is not completely understood, but it is considered that sunlight, genetic predisposition, pregnancy and hormonal therapy including oral contraceptives play more of a role (5-7).

It is helpful, before initiation of treatment, to perform an examination using wood’s lamp to recognize the depth of the melanin pigmentation and, thus, determine the type of melasma, including epidermal, dermal or mixed (8).

Despite the availability of various treatment modalities, treatment of melasma is often difficult and challenging (9).

Different treatment modalities including topical skin care regimens for suppressing melanin production (melanogenesis), chemical peels, micro-needling, mesotherapy, lasers and other light sources have been used with variable rate of success (10-15).

Q-switched (QS) Neodymium-doped Yttrium Aluminium garnet (Nd:YAG) lasers have been reported to be beneficial in some patients, but occasionally they cause complications like pigmentary change (hypo- or hyperpigmentation). In a study conducted by Kauvar in 2012, a new noninvasive combination treatment approach, using a low-fluence Q-switched Nd:YAG laser in conjunction with microdermabrasion was examined and found to be effective in treating melasma (1), but there was no control group in that study.

The purpose of the current study is to determine if this novel method is able to produce a significant improvement of melasma when compared with Q-switched Nd:YAG laser alone.

## Materials and methods

### Study design

We conducted a single-center prospective randomized clinical trial study of adults (>= 18 years of age) with split-face design. Patients diagnosed with moderate to severe melasma clinically, were enrolled.

20 patients (19 women and 1 man, aged 26-50, mean age: 34.2±7.3 years) with melasma lesions were included in this study. Inclusion criteria was patients older than 18 years with bilateral moderate to severe melasma. Exclusion criteria included pregnancy, breast feeding, Fitzpatrick skin types IV-VI, topical bleaching agent usage within 3 months before recruitment, employment of chemical peels, microdermabrasion or facial laser treatment within 9 months of entry, underlying skin disorders, taking isotretinoin or contraceptive pills during the past six months, sunlight or UV exposures, worsening of lesions or occurrence of post inflammatory hyperpigmentation (PIH) during the study.

The presence of moderate to severe melasma was based on a previously described melasma severity scale (16).

All patients were informed of the benefits and risks of the study after which they signed an informed consent form. The study was approved by local ethics committee (approval ID: IR.RUMS.REC.1394.279) and registered as IRCT20191228045917N2 in Iranian Registry of clinical Trials.

### Treatment protocol

20 individuals who met the eligibility criteria of the trial were subjected to a detailed review of their clinical history (age, sex, previous treatments, site of involvement) and physical examination. After this initial visit, all patients were given 4% hydroquinone cream (lightening cream Medillan®, Golafshan-e-arayesh, Iran) and adapalen gel (Acnopal®, Caspiantamin, Iran) to apply every other night to both sides of the face for a period of 4 weeks according to previous studies (1,17) and were told to stop applying them one night before each treatment session.

One side of the face randomly determined for treating with microdermabrasion by sealed envelopes numbered from 1 to 20 in which the allocation was indicated by simple randomization block. The envelopes were opened in an ascending order.

The side allocated to receive microdermabrasion prior to laser therapy, was treated with a total of 3 pass in each session. Microdermabrasion was performed using a standard protocol reported previously (18). The patients washed their face with daily facial cleanser(dry skin pain, Medipain®, Golafshan-e-arayesh, Iran).After degreasing with alcohol and waiting for the skin to dry, 3 pass microdermabrasion was done using 6 mm head and 400 mmHg pressure. The face was then rinsed with water and dried and immediately both sides of patient’s face was treated with Q-switched Nd:YAG laser (COSJET®, WONTECH, South Korea) with these parameters : wave length :1064 nm, fluence : 1.6 - 4 J/cm^2^ with a 4 or 6 mm spot size, using only 2 laser passes to achieve complete coverage of the affected skin. Visible frost or mild erythema of skin was taken as the end point.

At first session, fluence was set at 2J/cm2 for skin type II, 1.8 J/cm2 for skin type III and 1.6 J/cm2 for skin type IV with a 6 mm spot size. On next sessions, fluence was decreased or increased by 0.5-1 J/cm2 according to presence of side effects as well as patient tolerance. We achieved our desired results with fluences ranged from 1.6 to 4 J/cm2.

After laser therapy, patient face was cooled down using icepack and was covered with zinc oxide and fluocinolone cream. Patients were asked to apply a repair cream (Therapeutic cream Dr Jila®, Iran Avandfar, Iran) and also a sun-blocker cream (Sunscreen cream, oil free, SPF 60, Dr Jila®, Iran Avandfar, Iran) for 3 days before starting their hydroquinone cream and adapalen gel.

The participants were treated three times at regular time intervals of 4 weeks. All patients were examined prior to each treatment session and also 1 and 3 month after last session. All of visits were performed by one dermatologist.

On each visit, Melasma Area and Severity Index (MASI) as well as Patient Global melasma improvement Assessment (PGA) were determined and the patients were asked about any unfavorable side effect such as burning sensation, erythema, edema, blistering, erosion or crust formation and facial asymmetry.

Patients were also asked about the improvement of melasma on each side of their face (PGA) on each visit rating that according to the following numerical scale (19):

-1: worsening 0: no change 1: slight improvement

2: moderate improvement 3: marked improvement 4: complete clearance

The blinded investigator calculated the MASI score that quantifies the extent of the lesion (A), the darkness (D) and the homogeneity (H) of the hyperpigmentation. Each of the 4 areas including forehead, perioral/chin, right and left malar regions was scored. The sum of the severity rating for the darkness and the homogeneity is multiplied by the numeric value of the involved area to calculate the MASI score (range : 0 -48) (20,21), but in this study, MASI score was calculated for each side of face separately (range : 0 -24) with dividing the numeric value of the forehead and perioral / chin by 2.

### Statistical analysis

Statistical analysis were undertaken by using SPSS, version 23. P value of 0.05 or less was considered significant. All of the statistical analyses have been done with 95% confidence interval. Chi-square and Mc Nemar’s tests have been used to show correlations.

## Results

A total of 20 patients were enrolled in this study out of which 18 patients completed the study with no protocol deviations. The 2 subjects who did not complete the study were unable to come for last laser session or follow up sessions because of scheduling difficulties.

The majority of patients were female (95%) and had Fitzpatrick skin type III (80%). Mean age of participants was 34.2±7.3 years ranging from 26 to50.

Specific patient demographics is included in table 1.

**Table 1.**
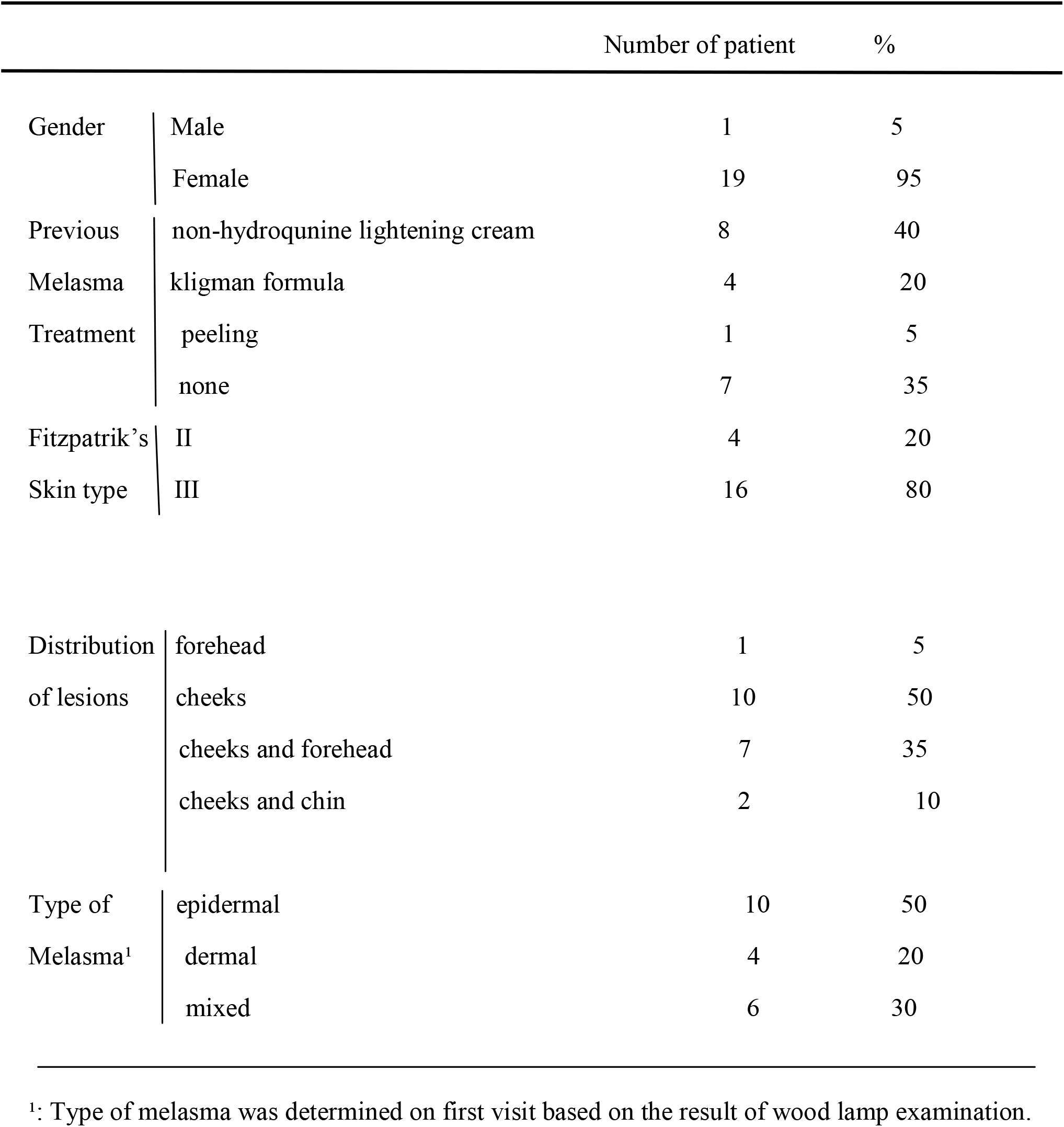
A summary of baseline characteristics of patients.

Mean baseline MASI score was 10.3 ±2.13 on the abraded side and 9.8 ±2.32 on the other side (Table2).

**Table 2.**
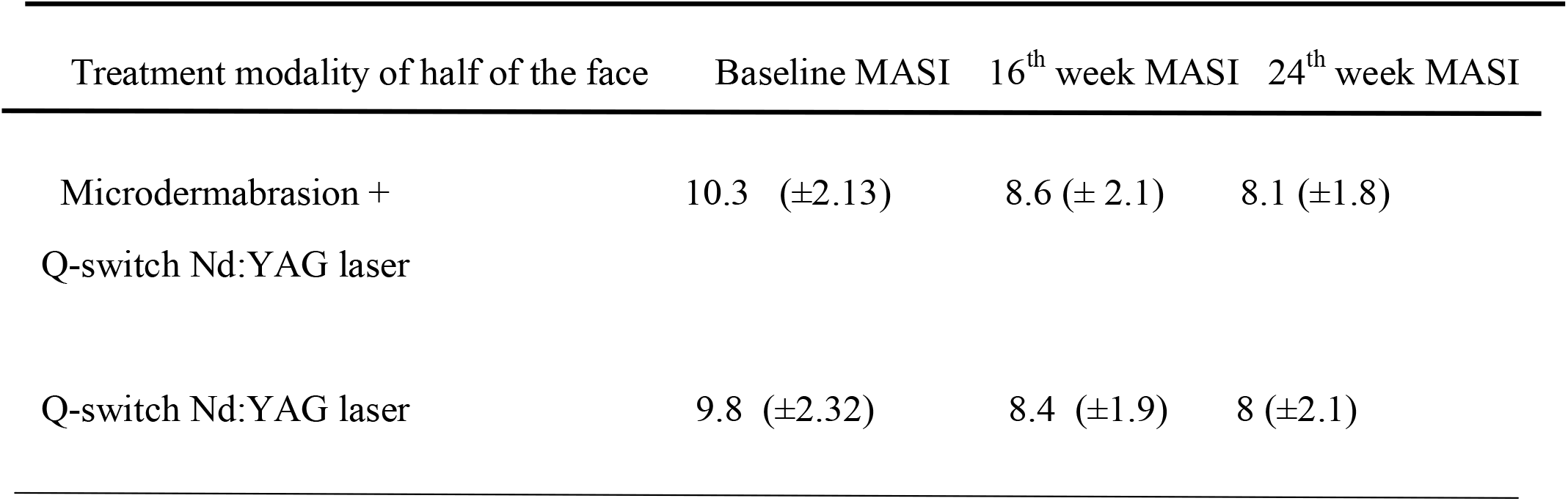
Means and standard deviations for MASI score on each side of face at baseline and follow up sessions.

Figure 1 shows the improvement in MASI score on both sides of the face. The baseline covariate for MASI was significant (P value < 0.001).

**Figure 1:**
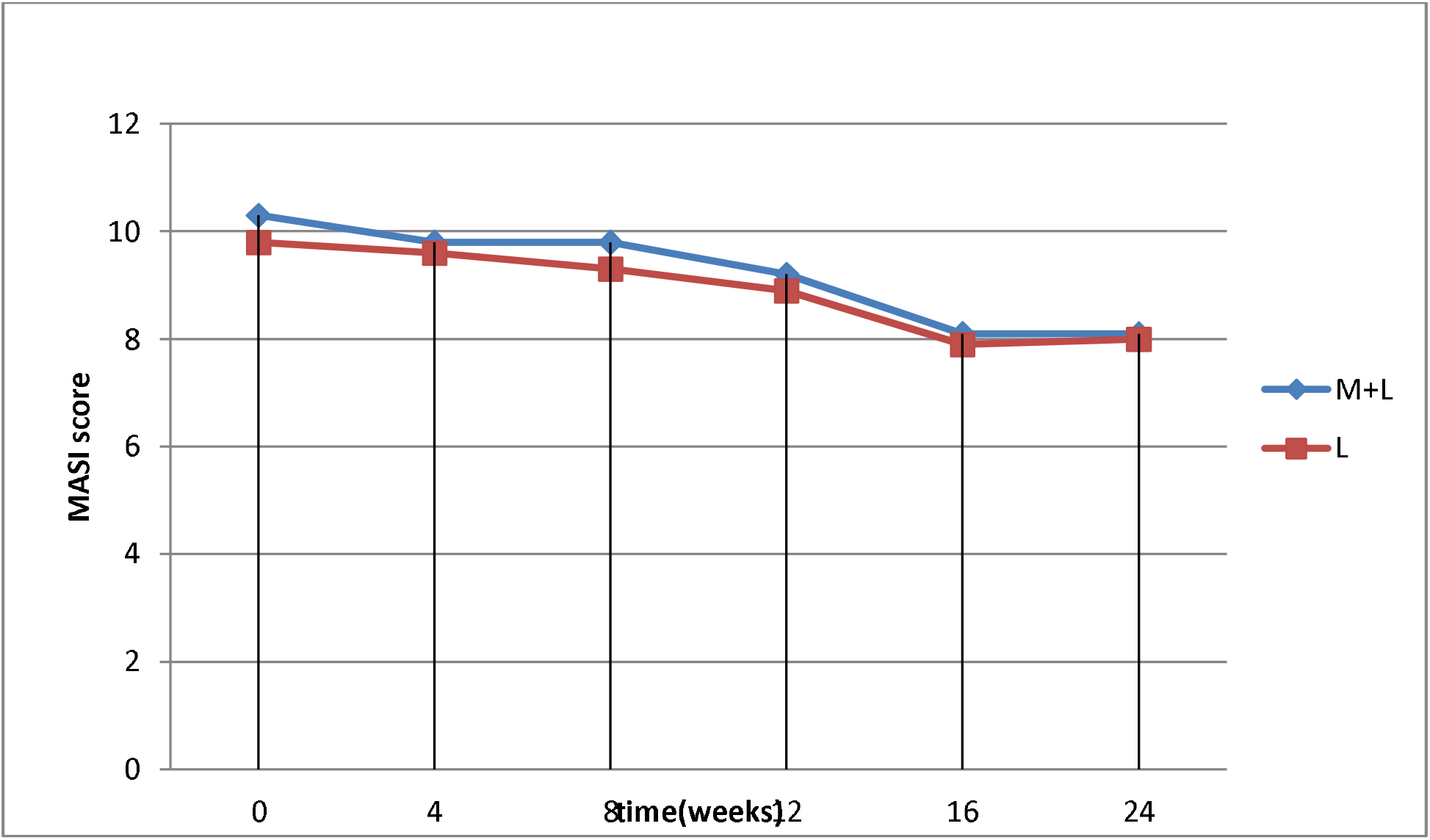
MASI score on abraded versus unabraded side. Abraded group effect: (abraded vs unabraded) P = 0.609. overall time effect: P < 0.001. (M+L: microdermabrasion and laser, L: only laser)

It is worthy to note that although both sides improved from baseline to the 24^th^ week (P value < 0.001), there was no statistically significant difference between both sides on 24^th^ week (Figure1).

The PGA at second follow up visit on 24^th^ week was 2.6 for abraded side and 2.2 for other side. There was not any statistically significant difference between two sides on 24^th^ week (P value: 0.33), but difference between PGA at 4^th^ week in comparison with PGA at 24^th^ week was meaningful for both sides (Table3).

**Table 3.** Mean value of PGA (Patient Global melasma improvement Assessment) index on both sides of face

| PGA | week 4 | week 24 | P value |
| --- | --- | --- | --- |
| for each half of the face | (baseline) |  |  |
| Laser with microdermabrasion | 1.6 | 2.6 | 0.04 |
| Laser only | 1.3 | 2.2 | 0.01 |

Adverse effects for both sides were found to be equal and consisted of erythema (60%) and burning sensation (50%) both lasting only for 1 to 2 days. Moderate facial edema lasting up to 4 days was reported by 10% of patients. Blistering, scaring, pigmentary changes and an asymmetric look during or after study were not observed.

## Discussion

In this study, 20 patients were treated with Nd:YAG Q-switched laser and microdermabrasion for melasma. Outcome measures selected for this study were subjective and interpreted by both physician and patient and a global evaluation of improvement as well as scoring index (MASI) have been used to help quantify the evaluation.

Using these evaluation methods, both sides of the face in our patients improved remarkably, but there was not any significant difference between them.

This study confirms the efficacy and safety of Nd:YAG Q-switched laser alone or with microdermabrasion in treatment of melasma.

Nd:YAG Q-switched laser has been found to be effective in treating melasma in several studies (22-27), with 67% of patients with moderate to severe disease achieving complete or near-complete clearance after 10 weeks of treatment (22).

Kunachak et al in a large-scale study of 533 patients with melasma, found that 97% of patients achieved clearance of melasma with dermabrasion (28), but complications such as keloid formation, milia, post inflammatory hyperpigmentation and loss of skin texture were seen. So dermabrasion is not a standard treatment modality. A noninvasive and minimal form of it, microdermabrasion, had been used in a study by Kauvar who investigated the efficacy of combination of microdermabrasion and low-fluence Q-switched laser in 27 patients with skin photo-type of II to V. All subjects achieved at least a 50% improvement in the appearance of their melasma. More than 80% of the subjects experienced >76% improvement in melasma at follow –up. However, that study lacked the control group (1).

According to some authors, combination treatment strategies may provide a more robust clinical outcomes in melsma treatment.

Theoretically, microdermabrasion might decrease scattering of laser light and lead to an increase in the depth of penetration of the Q-switched Nd:YAG laser beam. On the other hand, it can help with elimination of epidermal melanin by increasing epidermal skin turn over and exfoliating the stratum corneum. Moreover, it can improve lightening drugs penetration by exfoliating the most superficial layers of stratum corneum. Hence, it seems that microdermabrasion might add to the effect of Q-switched Nd:YAG laser in treating melasma.

We evaluated this hypothesis by using combination therapy versus Q-switched Nd:YAG laser alone in this split-face design study, but contrary to Kauver, we failed to report any difference between these two treatment modalities. This incongruity of results necessitates performing further studies with different methodologies and a larger number of patients to shed more light on this matter. As far as we know, current study is the first prospective randomized clinical trial evaluated efficacy and safety of this new combination. Therefore, we could not compare our results with previous studies. However, both treatment modalities were used previously and their clinical efficacy was found to be superior compared to topical agents (29-30) This study confirmed the efficacy and safety of laser with or without microdermabrasion, but we could not find any difference between two study groups in terms of clinical response or safety profile. In fact, none of our patients experienced post inflammatory hyperpigmentation which could be due to use of lightening agents before initiation of and during the study as well as using conservative setting for laser treatment. It should be borne in mind that a large part of this study was conducted during autumn and winter, which can be an important factor in a reduction in the incidence of sun exposure related side effects. In addition, another explanation for absence of any PIH or recurrence of melasma in our patients might be the fact that our follow up period was not long enough to assess the stability and durability of treatment. Consequently, different results might be expected from future studies with longer follow-up duration.

Despite the success of this study, there were some limitations must be considered including lack of devices for objective measures of clinical improvement as well as absence of histopathological evaluation due to disinclination of majority of patients for undergoing biopsy.

Using modern modalities for assessment of any change in lesion’s color and melanin content would provide less biased and more reliable results regarding clinical efficacy of both treatment modalities and is highly recommended in future studies.

Another limitation of this study was recruiting a small number of patients due to our strict recruitment criteria and the fact that in the city in which study was conducted, the majority of patients had skin photo types IV and higher which made the laser therapy unsuitable for them. The limited number of patients may account for the lack of statistically significant differences between the two groups and also led to inability to perform further subgroup analyses regarding demographics and clinical characteristics of patients. Moreover, even though we followed our patients for 3 months that seems to be a suitable time duration according to previous reports [31], longer follow up periods may lead to a better assessment of this treatment modality as mentioned before.

In conclusion, We have shown that combination therapy with Q-switched Nd:YAG laser and microdermabrasion is as effective as Q-switched Nd:YAG laser alone and does not add to the effect of laser in treating melasma.

Considering our study limitations, further prospective studies with a large number of patients and a longer follow-up period for validating our results are encouraged.

## Data Availability

The data that support the findings of this study are available on request from the corresponding author

## Acknowledgments

The authors thank Iran’s National Elites Foundation for providing financial support.

